# Feeling at Home in a Virtually Amputated Body; Neural and Phenomenological Effects of Illusory Embodiment in Body Integrity Dysphoria

**DOI:** 10.1101/2024.01.26.24301812

**Authors:** Gianluca Saetta, Yannik Peter, Kathy Ruddy, Jasmine T. Ho, Roger Luechinger, Emily Cross, Lars Michels, Bigna Lenggenhager

**Author notes:** Corresponding author: Gianluca Saetta, ETH Zurich, Professorship for Social Brain Sciences (SBS), Stampfenbachstrasse 69, STB G 13.1, CH-8092 Zurich.

## Abstract

In Body Integrity Dysphoria (BID) a profound incongruity between the physical body and the desired, i.e., amputated body, often leads to a desire for limb amputation. Virtual reality (VR) and multisensory stimulation paradigms provide powerful tools to create the experience of being embodied in an amputated body.

Here we investigate the impact of such an experience on neural and subjective responses in 18 individuals with BID and 18 controls. We used both task-based and resting-state MRI before and after participants played an immersive virtual game in an amputated body corresponding to their desired bodily shape and mimicking their movements. The task-based fMRI assessed neural activity when viewing images of the body in the desired versus the undesired state.

Individuals with BID reported higher sense of ownership and control over the virtual body. Task-based fMRI showed increased pre-VR activity in the right superior parietal lobule (rSPL), right angular gyrus, and right supplementary motor area in the BID group, normalizing after VR exposure. Resting-state fMRI showed reduced connectivity in the rSPL, visuo-occipital areas, fronto-parietal, and fronto-striatal mirror and limb system networks, also normalising post-VR. Additionally, there was a normalization in the pattern of increased connectivity of cortico-striatal tracts connecting the rSPL and the pars orbitalis of the right inferior frontal gyrus with the nucleus accumbens.

Our findings suggest that virtual embodiment effectively modulates BID-related neural networks, offering a safe, cost-effective intervention for BID and highlights VR’s potential in exploring the complex interaction between body and self, with potential implications for similar psychiatric conditions.

## Introduction

Individuals with BID experience a profound discontentment regarding their bodily configuration or functionality. In the amputation variant of BID, people identify with an amputated body despite a complete, fully-functional body. The prospect of an amputation, paradoxically fosters a sense of completeness (1). Previous neuroimaging findings highlight atypical structures and functions of the rSPL related to this condition (2–5). The rSPL, a key substrate for body image(6), shows reduced gray matter and decreased functional and structural connectivity compared to healthy controls. The reduced connectivity, especially with the visual parieto-occipital area, might underlie the mismatch between the internally represented body image of a missing limb and the visually perceived “fully-limbed body” (7, 8).

BID, recognized in the last revision of the International Classification of Diseases (ICD-11), arises from complex neurological, psychological, and social factors (9). The life quality is influenced by the time spent simulating their desired body state, e.g. by using crutches or wheelchairs, often referred to as “pretending” (10). The behaviour is a coping mechanism to ease the incongruence between the current and desired body state, thereby alleviating associated distress (10, 11). Yet, excessive simulation can result in both short- and long-term adverse effects, like compromised blood circulation (8). A potential alternative and safer approach could involve the application of brain-computer interfaces (12) or embodied virtual reality (VR). The latter enables the experience of owning and controlling from a first-person perspective avatars of diverse body shapes or functionalities through multisensory contingencies. Participants can embody avatars of different body size (13) or gender (14, 15), altering in a broad range of physiological perceptual, cognitive, emotional, and social processes, and therefore showing great therapeutic potential (16). Here, we use VR to enable embodiment of the BID individual’s desired (i.e. amputated) body. A preliminary study with two individuals with BID demonstrated a decrease in BID symptoms after experiencing a virtually amputated body as their own (17). We investigate the effect of a playful, immersive VR game played from a first-person perspective in an individually amputated avatar that follows the participant’s movements (sensorimotor contingency) on neural and subjective measures in a group of 18 individuals with BID and a matched control group.

## Materials and Methods

### Participants

The study cohort consisted of 18 Individuals with BID, encompassing 5 women and 13 men (mean age = 45.83 ± 14.00 years). Additionally, a group of 18 healthy age matched (Welch two-sample t-test on age: t(35) = −0.52, p = 0.61) controls, comprising 4 women and 14 men (mean age = 47.83 ± 13.92 years), was included. One participant from the BID group experienced discomfort during the MRI session and thus provided only the phenomenological data. Ten individuals with BID reported a desire for amputation of their left leg, while 8 desired amputations of their right leg. Two participants had already participated in a previous neuroimaging study (7, 8))

Five Individuals with BID but none of the healthy controls reported Gender Identity Dysphoria (3 biologically male identifying as female (i.e., male-to-female transgender), one biologically female identifying as male (i.e., female-to-male transgender), and one biologically male who previously did a gender reassignment surgery, which is in line with previous literature showing a comorbidities between the two conditions (27).

Individuals with BID were recruited through mailing lists, online self-help groups and personal contacts. As BID is a relatively rare condition, these individuals were recruited from different countries (Germany: n = 7; Switzerland: n = 5; USA: n = 2; UK: n = 2; Netherlands: n = 1; Austria: n = 1).

The evaluation of BID diagnostic criteria was performed by trained psychologists according to the inclusion criteria provided by the ICD-11. The intensity of the desire for amputation was assessed through the Zurich Xenomelia Scale.

Eligibility criteria included: (i) age range (18-65 years), (ii) provided informed consent, (iii) gender diversity (male, female, non-binary), and self-reported BID with a desire for limb amputation (for the BID group, or lack thereof for the control group). Exclusion criteria were: (i) presence of major psychiatric or neurological disorders beyond BID, including depression and anxiety; (ii) non-compliance with study protocols; (iii) evidence of drug or alcohol abuse, as determined through self-report measures; (iv) inability to adhere to study procedures due to language barriers or underlying psychological conditions; (v) contraindications for MRI examinations based on questionnaire screening. The study was approved by the local Ethics Committee of the University Hospital of Zurich (EK-BASEC: ID 2021-01867) and complied with the Declaration of Helsinki (1964).

### Study Procedure

To assess the clinical features of a desired amputation in BID and their potential modification through embodied VR interventions, individuals with BID engaged in an online survey administered via the Psytoolkit software (45) before and after the main study. Each survey lasted approximately 10 minutes. On the day of testing, both groups underwent MRI scans, repeated before and after the VR intervention: structural MRI scans, rs-fMRI, and task-based fMRI. In the post-VR intervention session, Diffusion Tensor Imaging (DTI) data, to be discussed separately, were collected. Data collection took place at the Institute of Biomedicine (ETH Zurich). After each MRI session, participants rated the images encountered during the active task fMRI. For the sequence of task administration see Fig. 1.

**Figure 1.**
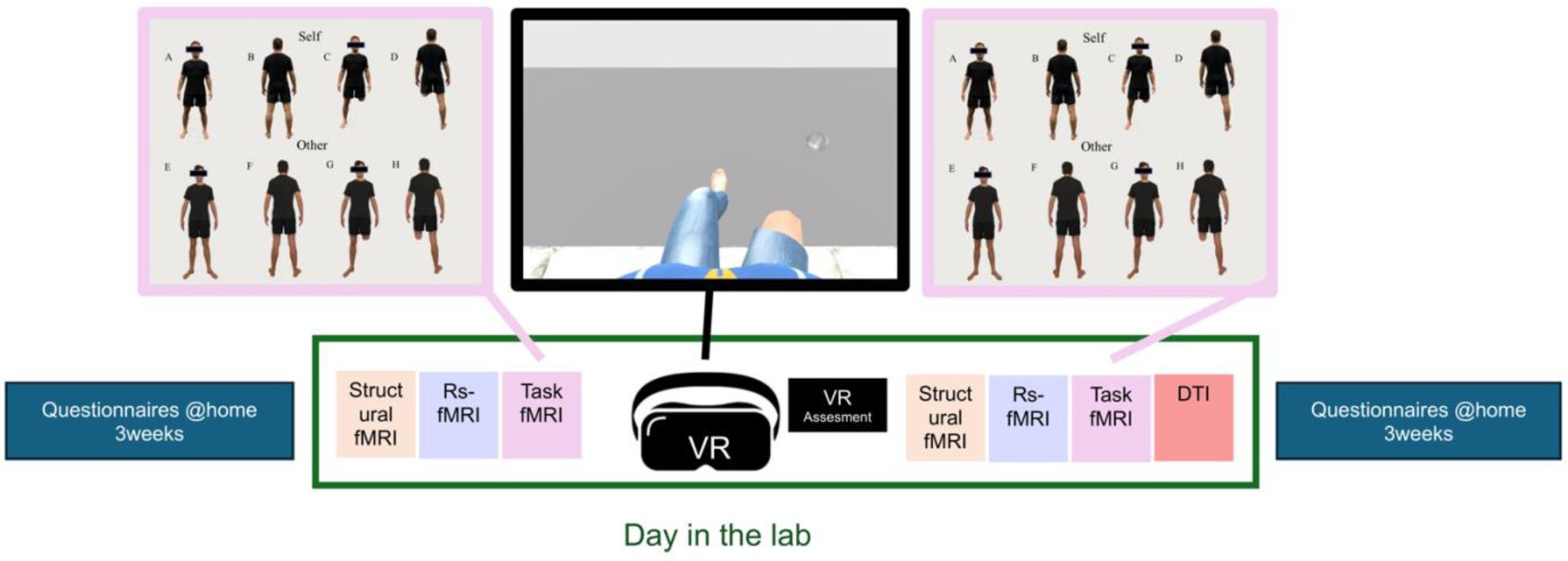
Study Procedure and Task-based fMRI Experiment. BID participants, but not controls, completed online clinical assessments once a week for 3 weeks before and after the lab visit. For BID and control groups Lab tasks included in the following order: (i) T1-weighted morphological data; (ii) rs-fMRI; (iii) task-based fMRI; (v) VR experience; (vi) VR Embodiment assessment (vii) rs-fMRI; (viii) task-based fMRI; (ix) DTI data (not discussed here). Task-based fMRI conditions were: A-D: Self, E-H: Other, with variations in view and amputation. VR used a first-person perspective. Leg movements were tracked and displayed on the avatar. The task involved exploring their body or catching soap bubbles with the virtual stump.

### Data Collection and Analysis

While raw data will be made available in a separate data descriptor contribution, processed behavioral data and relative scripts as well as the MATLAB (spm.mat) file to explore the neuroimaging findings are available on Open Science Framework (OSF - https://osf.io/qag6u/)

### Virtual embodiment

A head-mounted display (HMD, Oculus Rift S) was used for visual stimulation in VR. The Oculus Rift S employs a rapid-switch LCD panel with a display resolution of 1280×1440 pixels per eye, operating at an 80 Hz refresh rate. To facilitate tracking of head and controller positions, the integrated cameras of the Oculus Rift S were utilized. This tracking data was integrated into Unity Version 2020.3.12f1 to animate avatars crafted using the open-source tool MakeHuman. Subsequent avatar refinement took place in Blender Version 2.93.11, creating a series of avatars with customized amputations at locations defined by participants. The avatars matched the desired amputation in size and height for individuals with BID. Healthy controls, matched for age, gender, height, and side of amputation with the individuals with BID, underwent the same virtual amputation procedure as their BID counterparts.

Given the three-point tracking data supplied by the Oculus Rift S (head, left controller, right controller), animations were required to simulate leg movements. In this regard, inverse kinematics were applied using the Animation Rigging Toolbox Version 1.0.3, coupled with positional tracking data from the two controllers, with one attached to each of the participants’ feet.

During VR stimulation, participants sat on an examination table with their legs hanging off one side. The HMD was worn, with controllers attached to the feet. The virtual avatar’s position and dimensions were adjusted within the VR environment until participants perceived a natural appearance and size. Manual adjustments were made to the hip position based on participant posture. Additionally, slight size adjustments were made for participants with differing leg lengths. Participants embodied a virtual avatar with a leg amputation from a first-person perspective with an individually defined line of demarcation between accepted and rejected body segments. For the affected leg, a stump was visible (see Fig. 1, in the desired state for BID but the undesired state for controls; videos are available on OSF (https://osf.io/qag6u/)).

The stimulation protocol encompassed eight rounds, each lasting 120 seconds. Participants engaged in one of two tasks during each round: exploring the avatar by observing and moving its legs or participating in a game involving popping soap bubbles with the virtual stump. The sequence alternated between exploration and gameplay, repeated four times for a total of eight rounds, requiring 16 minutes.

Following the completion of these rounds, participants answered a seven-item questionnaire on embodiment (in English or German) via a visual analogue scale (VAS) from 0 to 100. They used head movements to control a cursor, indicating their responses on a bar without seeing the selected value. The questions addressed five conditions: *Virtual Body Ownership*, *Virtual Body Agency*, *Virtual Leg Ownership*, and *Virtual Leg Agency*. The exact questions are reported in the Supplementary Information (SI).

At the end of the VR experience, before starting the post-VR MRI session, individuals with BID were asked if they experienced an increased desire for amputation.

Data analyses were performed using R Studio v. 1.3.1093, with packages coin and ARTool. An alpha level of 0.05 was set for significance. Normality was tested with a Shapiro-Wilk test and visual inspection of residuals. Non-parametric tests were used accordingly. Wilcoxon rank sum tests assessed differences in VAS scores between the BID and control groups for *Virtual Body Ownership*, *Virtual Body Agency*.

For *Virtual Leg Ownership* and *Virtual Leg Agency*, we created a 2-level factor, *Virtual Amputation* (yes/no). A non-parametric ANOVA (ART-ANOVA) was calculated, considering the between-factor *Group* (BID/Control), the within-factor *Consistency Amputation* (consistent/non-consistent), and their interaction. Post-hoc tests were performed using the “art.con” function.

### Task-based fMRI

The fMRI experiment used a block-designed approach with four functional runs, following previously outlined parameters and task design (23). The stimulus set featured participant images, edited with GIMP software to represent the participants in their desired body state (e.g., with a leg amputation for the BID group) and their physical state (complete for the BID group), presented from front and back angles. Four stimulus categories were defined: “Self vs Other” and “*Full vs Amputated Body*.” In the “*Other*” category, age- and sex-matched individuals had their legs digitally removed to simulate the desired limb amputation characteristic of BID (see Fig. 1). Each condition included eight stimuli, presented three times, totalling 24 trials. Rest intervals lasting 9-12 seconds, with an average jittered mean of 10.5 seconds interspersed these trials.

Participants were instructed to attentively observe the presented images and register any emotional responses, following the methodology described previously (23). Serving as an attentional control task, each run concluded with images of a green square (n = 5) or a pink square (n = 5). Participants promptly responded by pressing a handheld button using either their index or middle finger, corresponding to one of the squares. Analysis and results will be presented in a separate data descriptor paper.

Participants were scanned using a 3.0 Tesla Philips Ingenia whole-body scanner (Philips Medical Systems, Best, The Netherlands). Acquisition parameters and preprocessing steps which followed a standard procedure are described in the SI.

A fixed-effects analysis was conducted for each subject to characterize the BOLD response associated with each picture block relative to its baseline condition. The contrast images from this analysis were entered into a random-effects second-level group analysis. A full factorial analysis was performed, considering *Group* (BID vs. Controls), *Session* (Pre vs. Post VR), and *Body State* (Full vs. Amputated). The design matrix had 8 columns. Importantly, whole-brain analysis without masking was applied. We exclusively report predefined hypothesized effects using the most conservative thresholding method available in SPM (p < 0.05 familywise (FEW) voxel-wise corrected) with a cluster-extent threshold of k > 25 to exclude spurious activations.

### Rs-fMRI data

We also collected rs-fMRI spin-echo echo-planar imaging (EPI) scans. Participants were instructed to relax while staring at a fixation cross displayed on a screen, allowing their minds to wander. Acquisitions parameters and preprocessing steps which followed a standard procedure are described in the SI.

After preprocessing and denoising, at first-level of analysis, ROI-to-ROI connectivity matrices were estimated characterizing the functional connectivity between each pair of regions among 28 Harvard-Oxford atlas ROIs (46), with a hypothesis-driven approach (7). The ROIs for the connectivity matrices include the left frontal pole (lFP), right frontal pole (rFP), right frontal orbital cortex (rFOrb), left frontal orbital cortex (lFOrb), right inferior frontal gyrus, pars triangularis (rIFGtri), left inferior frontal gyrus (lIFG), pars triangularis (lIFGtri), right inferior frontal gyrus, pars opercularis (rIFGoper), left inferior frontal gyrus, pars opercularis (lIFGoper), right precentral gyrus (rPreCG), left precentral gyrus (lPreCG), left middle frontal gyrus (lMidFG), left postcentral gyrus (lPostCG), rSPL, right angular gyrus (rAG), right lateral occipital cortex, superior division (rsLOC), right cuneus (rCuneal), posterior cingulate gyrus (PC), right middle temporal gyrus, anterior division (rMTG), right caudate (rCaudate), left caudate (lCaudate), right putamen (rPutamen), left putamen (lPutamen), right accumbens (rAccumbens), left accumbens (lAccumbens), right supplementary motor cortex (rSMA), and left supplementary motor cortex (lSMA).

Functional connectivity strength was estimated separately for each pair of ROIs characterizing the association between their BOLD signal timeseries. Functional connectivity strength was represented by Fisher-transformed bivariate correlation coefficients from a weighted general linear model (weighted-GLM, defined separately for each pair of seed and target areas, modelling the association between their BOLD signal time-series.

Individual scans were weighted by a boxcar signal characterizing each condition convolved with an SPM canonical hemodynamic response function and rectified.

Group level analyses were performed using a GLM. For each ROI-to-ROI connection, a separate GLM was estimated, with first-level connectivity measures at this connection as dependent variables and *Group* (BID vs Control) and *Session* (Pre vs Post) as independent variables. Connection-level hypotheses were evaluated using multivariate parametric statistics with random-effects across subjects and sample covariance estimation across multiple measurements. Inferences were performed at the level of individual clusters (groups of similar connections). Cluster-level inferences were based on parametric statistics within- and between-each pair of networks (Functional Network Connectivity (48), with networks identified using a complete-linkage hierarchical clustering procedure) based on ROI-to-ROI anatomical proximity and functional similarity metrics (49). Results were thresholded using a *p* < 0.05 connection-level threshold and a familywise corrected p-FDR < 0.05 cluster-level threshold (49).

### Supplementary Dataset for Replication Purposes

T1-weighted morphological data, Task-related picture rating outside of the scanner, clinical data and online survey in BID methods and Task-related picture rating outside of the scanner results described in the SI.

Investigation, Data curation, Project administration, Formal analysis, Writing – Original draft; Yannik Peter: Investigation, Data curation, Project administration, Formal analysis, Writing – Reviewing and Editing. Kathy Ruddy: Supervision, Writing – Reviewing and Editing; Jasmine Ho:, Methodology, Writing – Reviewing and Editing; Roger Luechinger: Investigation, Data curation, Writing – Reviewing and Editing; Emily Cross: Supervision, Lars Michels: Methodology, Project administration, Supervision, Writing – Reviewing and Editing; Bigna Lenggenhager: Funding acquisition, Conceptualization Methodology, Project administration, Supervision, Writing – Original Draft & Reviewing and Editing.

## Results

### Intensity of the desire for amputation in individuals with BID

The group means and standard deviations for each participant for the Zurich Xenomelia Scale, ranging from a minimum score of 4 indicating the lowest severity to a maximum score of 24 indicating the highest severity of symptoms, were as follows: *Pure Amputation Desir*e: 21.22 ± 3.13; *Erotic Attraction to amputated bodies*: 16.39 ± 5.07; *Pretending Behavior*: 17.00 ± 2.56

### Phenomenological experience during the embodied VR intervention

The Wilcoxon-Mann-Whitney tests were conducted to explore the relationship between Visual Analog Scale (VAS) scores and different groups across four specific conditions: *Virtual Body Ownership (*feeling the virtual body or the virtual leg is theirs), *Virtual Body Agency* (feeling they control the virtual body’s movements) and two conditions *Virtual Leg Ownership* and *Virtual Leg Agency* (ownership and agency inquired separately for the virtually amputated and intact leg)

For *Virtual Body Ownership*, BID showed higher VAS scores compared to controls (Z = 3.46, p = 0.001), indicating a heightened sense of ownership for the virtual body.

For *Virtual Body Agency*, the Asymptotic Wilcoxon-Mann-Whitney Test indicated no significant difference between the BID and control groups (Z = 1.78, p = 0.073).

For *Virtual Leg Ownership,* the ART-ANOVA, showed a statistically significant effect of *Group* (F = 15.84, p = 0.001), suggesting higher leg ownership ratings in BID compared to controls. The factor *Virtual Amputation* did not show a statistically significant effect (F = 0.32, p = 0.57). The interaction effect between *Group* and *Virtual Amputation* was statistically significant (F = 8.59, p = 0.006). Significant post-hoc tests (p < 0.05) indicate higher leg ownership ratings in BID for the amputated virtual leg *versus* Controls for the non-virtually amputated leg (Contrast: BID, virtually amputated vs Control, non-virtually amputated; Estimate = 8.528, SE = 3.36, t = 2.537, p = 0.016). Furthermore, leg ownership ratings notably increase in BID *versus* the Control, for the virtually amputated leg (Contrast: BID, Amputation Consistent vs Control, Amputation Consistent: Estimate = 17.528, SE = 7.52, t = 2.332, p = 0.026).

For *Virtual Leg Agency,* the ART-ANOVA, showed a statistically significant effect of *Group* (F = 6.00, p = 0.019, suggesting higher leg agency in BID compared to controls. The factor *Virtual Amputation* did not show a statistically significant effect (F = 2.71, p = 0.109). The interaction effect between *Group* and *Virtual Amputation* was not significant (F = 3.18, p = 0.083). All the results are displayed in Fig. 2.

**Figure 2.**
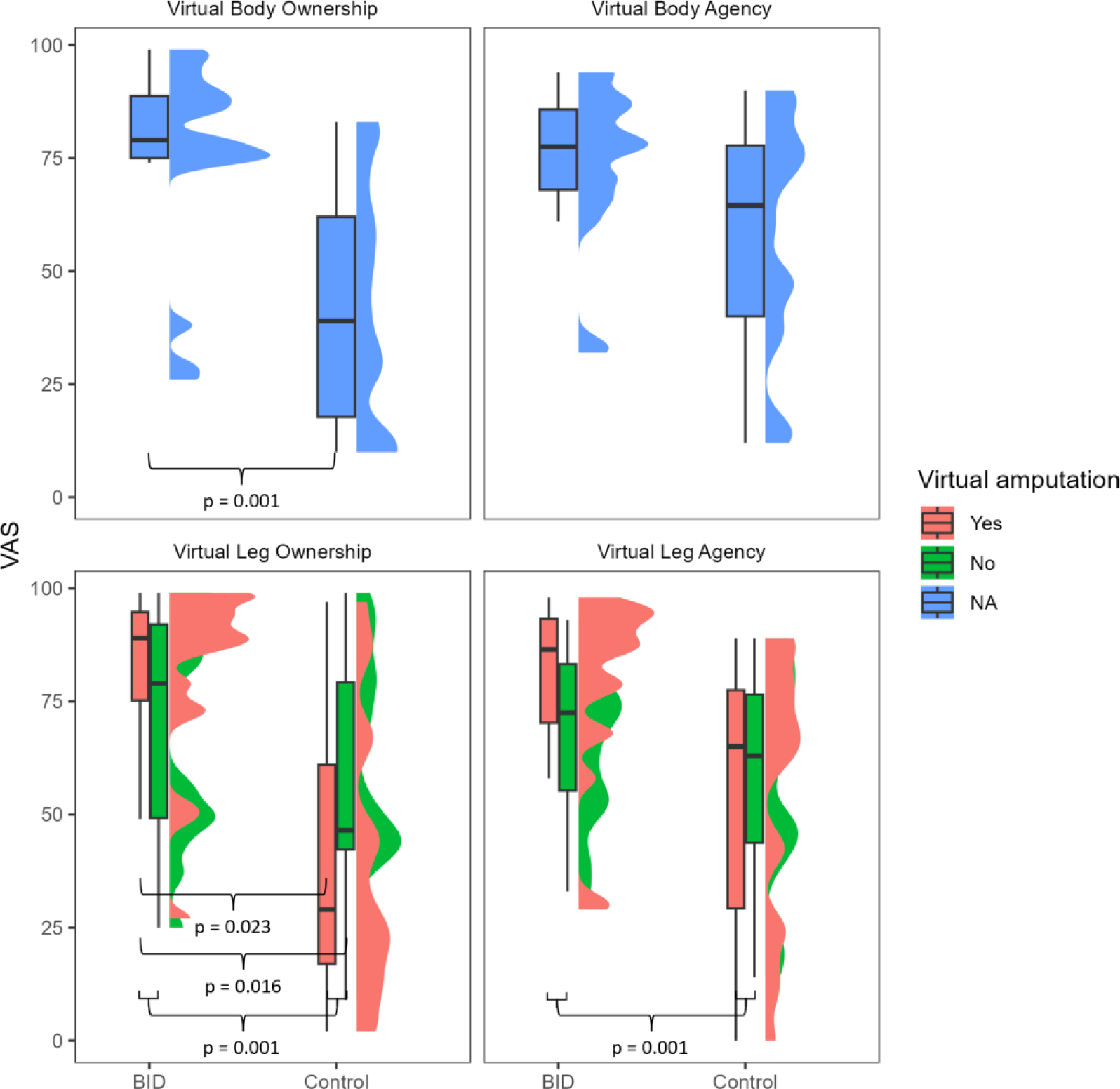
Phenomenological results of the virtual embodiment task. Boxplots and interquartile ranges are plotted.

Notably, in a debriefing performed after the VR *none of the individuals with BID* reported an increased desire and urge for amputation.

### Task-based fMRI

We specifically tested the a priori hypothesis that individuals with BID prior to exposure to VR, would display heightened activity in rSPL when exposed to images depicting their desired amputated state, in contrast to control subjects. This heightened activity, was expected to decrease post-VR treatment.

To test this hypothesis, upon observing a significant main effect of *Group* (refer to supplementary materials) and a non-significant effect of *Session*, we applied the contrast BID-PreVR-Amputated > BID-PostVR-Amputated. In line with our hypothesis, significant effects were observed in a cluster encompassing the rSPL and rAG.

Subsequently, effects were observed in the rSMA and another cluster positioned between the rPreCG and rPostCG (*p* < 0.05 FWE voxel-wise corrected, k > 25). The same contrast applied to the control group (Control-PreVR-Amputated > Control-PostVR-Amputated), along with contrasts utilizing opposite signs (BID-PreVR-Amputated < BID-PostVR-Amputated and Control-PreVR-Amputated < Control-PostVR-Amputated), produced non-significant results (*p* > 0.001 uncorrected). Detailed results can be found in Fig. 2A and Table 1A. Any additional main and interaction effects, which are not pertinent to our specific hypotheses, can be explored by accessing the spm.mat shared on the OSF (https://osf.io/qag6u/).

**Table 1.**
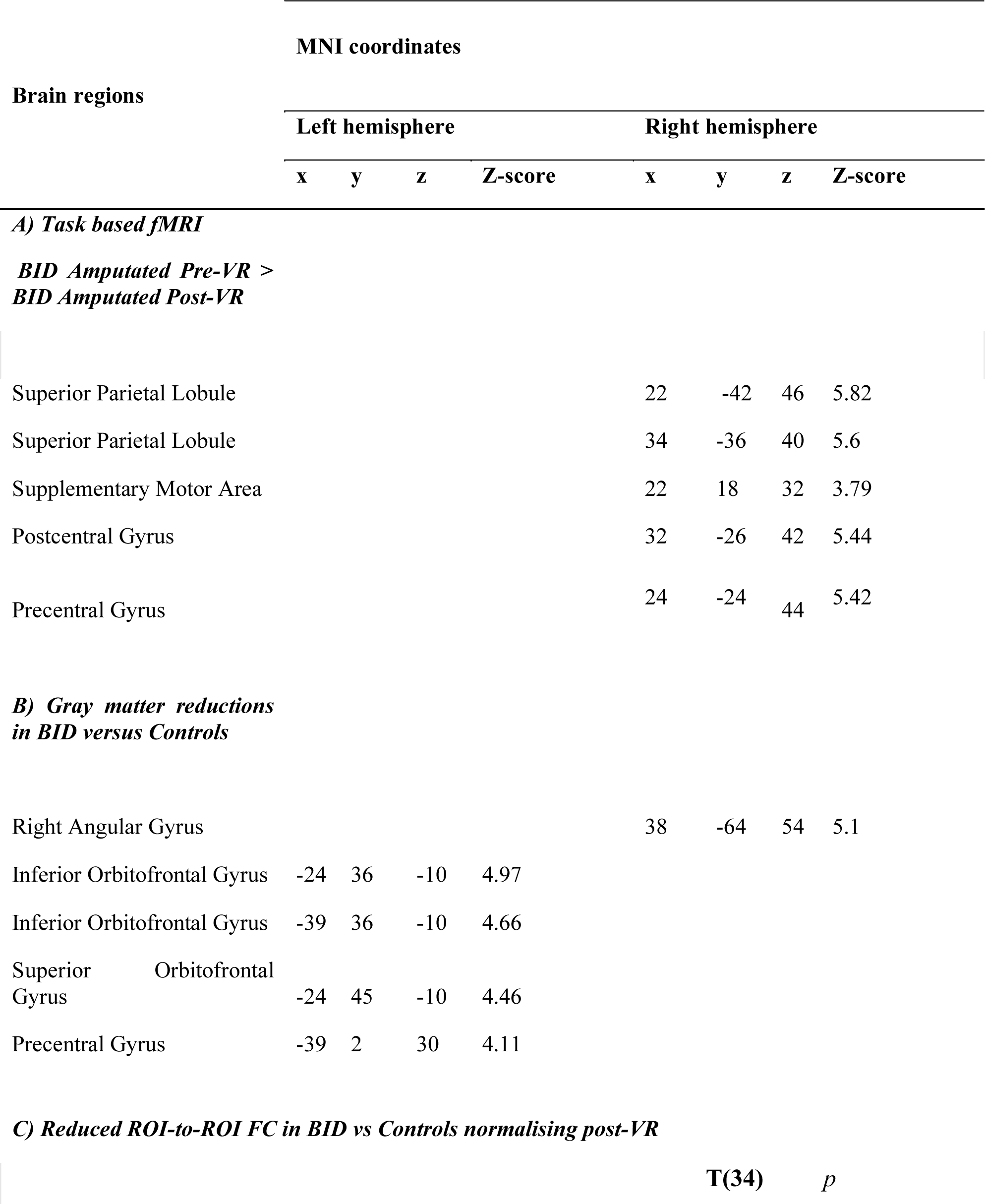
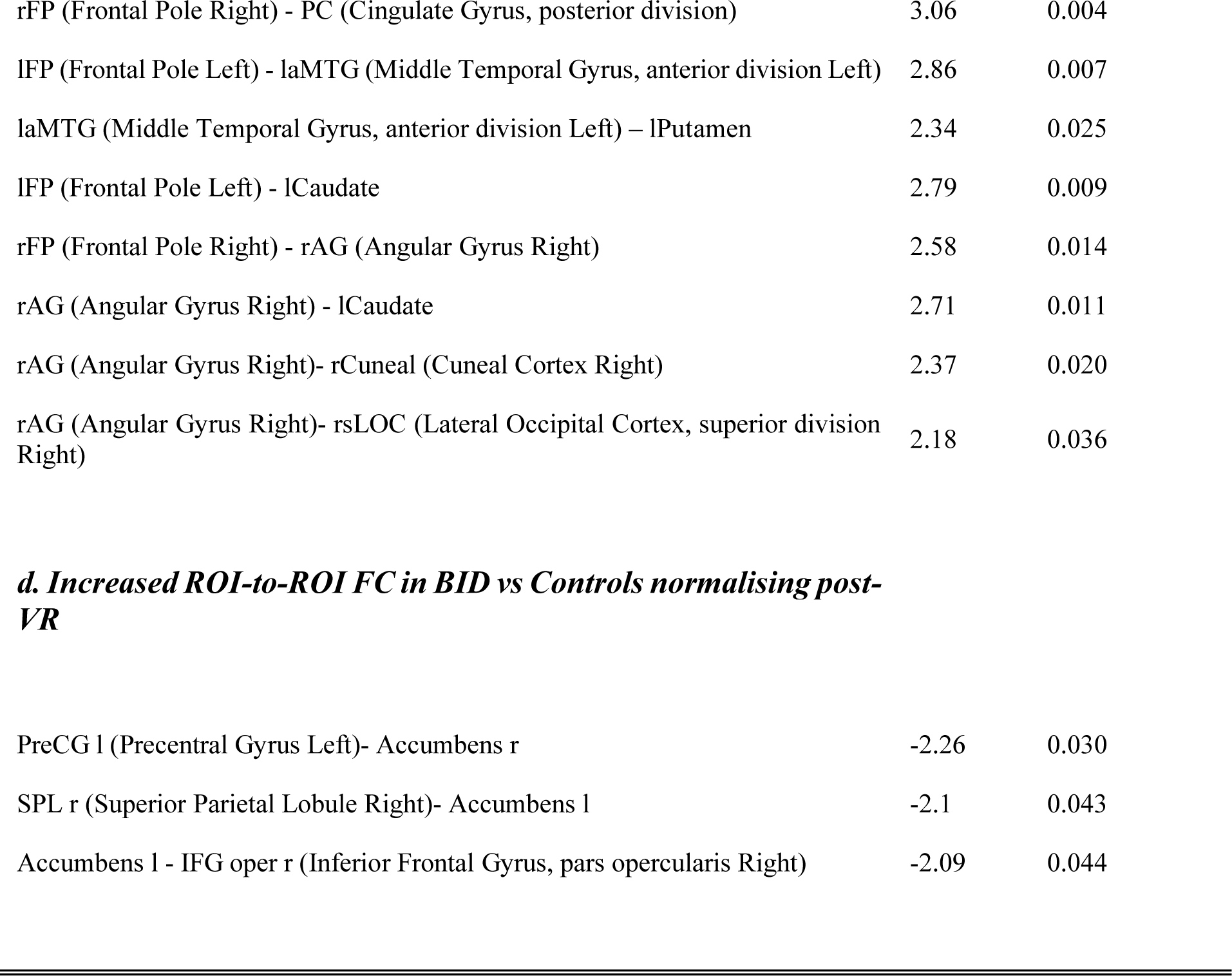
Result for task-based fMRI, T1-weighted morphological data and rs-state functional connectivity.

### Rs-fMRI

We hypothesized that the BID group would exhibit reduced ROI-to-ROI FC of the rSPL/rAG, visual parieto-occipital areas and areas within the limbic system in PreVR compared to Controls, that normalizes PostVR. We first observed the main effects of *Group* and *Session* (see supplementary materials). Consistent with this hypothesis, for whole brain analysis, the contrast Control > BID, PreVR > PostVR showed reduced ROI-to-ROI functional connectivity between the rAG and the following regions: the rCuneal, the rsLOC and the rCaudate. The rFP showed reduced connectivity with the rAG and the PC while the lFP showed reduced connectivity with the laMTG, and the lCaudate.

Additionally, the aMTG was less functionally connected to the lPutamen. Detailed results can be found in Fig. 2B and Table 1C. For PostVR data, the contrast BID > Control and Control > BID yielded no significant results, suggesting a normalization of those networks.

Also, in line with our hypothesis, the opposite contrast BID > Controls, PreVR > PostVR showed a normalization at PostVR of the enhanced reward-system connectivity that was observed PreVR. Specifically, this was between the lPreCG, the rAccumbens, the rSPL and the lAccumbens, and the rIFGoper and lAccumbens (see Fig. 2B and Table 1D). Any additional main and interaction effects can be explored by accessing the material shared on the OSF (https://osf.io/qag6u/).

### T1-weighted morphological data

For replication purposes, the analysis of the structural scans was expected to show lower gray matter volume in regions involved in the body image, multisensory integration, and the reward system. Consequently, the contrast was computed as Control > BID. Consistent with findings from prior research (Saetta et al., 2020) gray matter reductions in BID were observed in the rSPL, rAG, the left premotor cortex (in a cluster comprising the lPreCG and the lMidFG) and the pars orbitalis of the left inferior frontal gyrus (in a cluster comprising the lFP and lFOrb). Detailed results can be found in Fig. 2C and Table 1B. The spm.mat for further exploration of the results can be found on OSF (https://osf.io/qag6u/).

**Figure 2.**
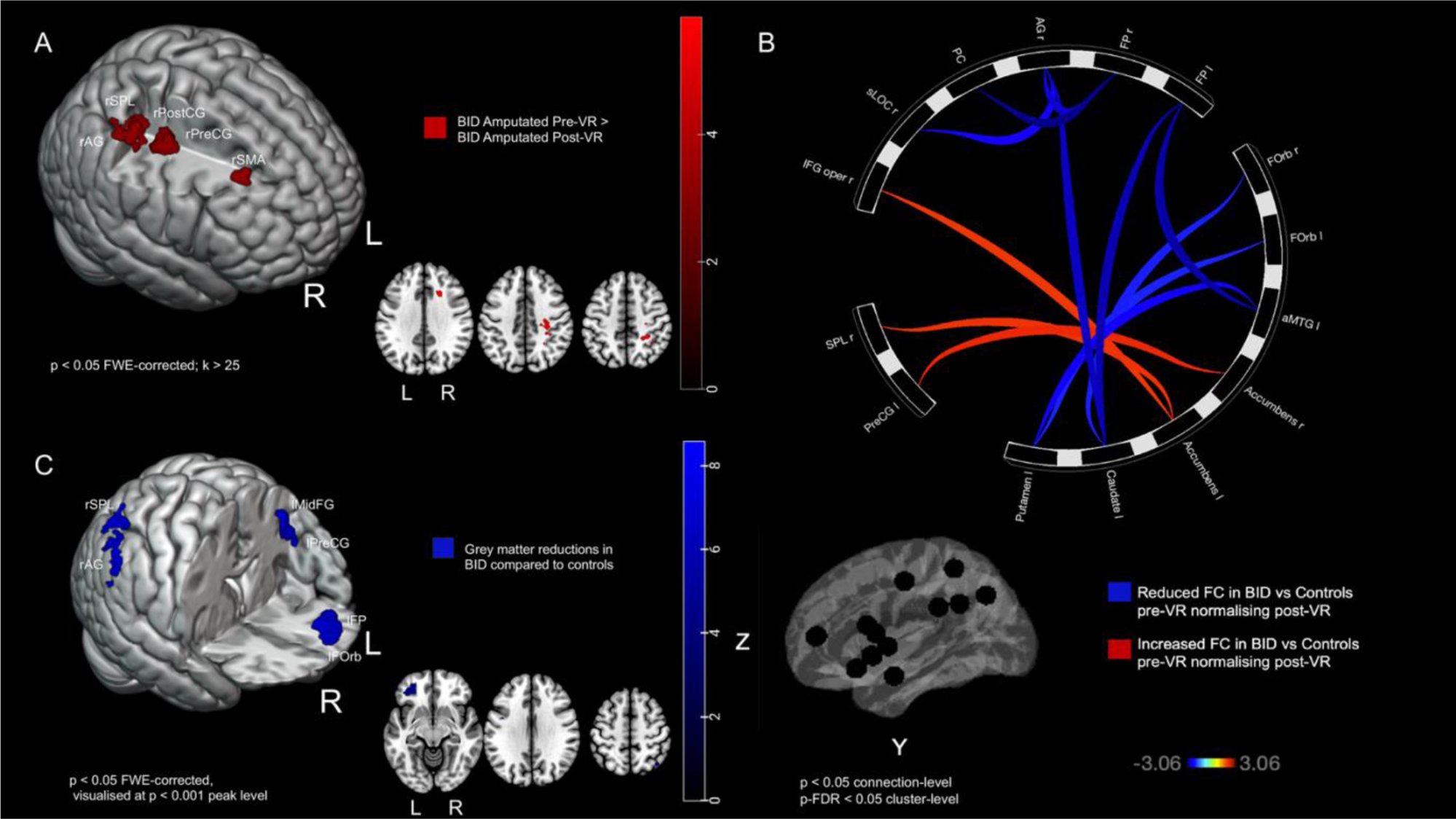
(f)MRI results in BID versus controls. A) Task-based fMRI results show enhanced neural patterns in BID participants before the VR intervention while viewing their desired body state, with normalization afterward. Key areas: right superior parietal lobule (rSPL), right angular gyrus (rAG), right postcentral gyrus (rPostCG), right precentral gyrus (rPreCG), and right supplementary motor area (rSMA). B) Resting-state fMRI results reveal reduced (blue) and increased (red) ROI connectivity in BID participants before the VR intervention, normalizing post-intervention. Abbreviations are detailed in the methods section of the main manuscript. C: Gray matter reductions in BID participants compared to controls are observed in the rSPL, rAG, left middle frontal gyrus (lMidFG), left precentral gyrus (lPreCG), left frontal orbital cortex (lFOrb), and left frontal pole (lFP).

## Discussion

In this study, we investigated how a short-term intervention based on embodiment of an amputated avatar influences neural and phenomenological underpinnings of BID. Individuals with BID often engage in pretending behaviors to align their physical body to their body image. We argue that VR offers a safer and more precise way to achieve this temporary alignment.

Both BID and control groups participated in a VR game, embodying an amputated avatar to catch soap bubbles using a virtual stump synchronized with their actual leg movements.

As expected, BID individuals reported higher ownership of the virtual amputated body compared to controls, aligning with previous findings of increased self-identification with an amputated body in BID (18, 19). Contrary to predictions, there was no significant difference in the sense of agency. While body ownership in BID is well-documented, the sense of agency is less explored. Recent research (20) suggests BID individuals have a reduced sense of agency for the affected limb, possibly explaining the smaller difference in agency for the virtual avatar. Our data show a relatively high sense of agency for the virtual body in both groups, suggesting a more holistic nature of the sense of agency in healthy participants that extends beyond specific body parts (21). BID individuals showed a higher sense of ownership, particularly for the virtual stump, while there was no group difference for the non-affected leg. In summary, BID individuals strongly embodied the amputated virtual avatar, reporting high ownership and agency.

The primary aim of the study was to investigate how virtual embodiment might affect neural activity and functional connectivity in previously identified BID-specific neural networks (7, 8). In task-based fMRI, the rSPL/rAG and rSMA showed enhanced activity before the VR when participants saw themselves in the desired state, which normalized after the VR exposure.

Task-based fMRI showed that rSPL/rAG and rSMA activity, enhanced before VR when participants saw their desired state, normalized after VR exposure. The rSPL integrates sensory visual, tactile, motor inputs to form a dynamic body image. Individuals with BID can perceive the “foreign” limb but fail to integrate it into their body image (5).

Consistently, in a recent study (22) BID individuals showed hypoactivity in the rSPL when the undesired leg was stimulated, indicating a failure to integrate the limb into their body image. Here in line with a previous reports (23), we observed heightened rSPL activity when BID individuals viewed their own amputated body, aligning with our hypothesis that the rSPL function as a *comparator* detecting “the lack of body adherence”, namely the discrepancies between the current body state and the body image. This activity normalized after VR, suggesting that VR embodiment can resolve these discrepancies.

The body image in the brain is believed to be innate and hardwired. This is demonstrated by sensations of limbs resembling the canonical form despite their physical absence from birth (e.g., amelia,(24) or unconventional forms, such as hands or feet attached directly to the trunk (e.g., phocomelia (25)). Further evidence includes cases of individuals experiencing a phantom penis and phantom erection after surgical removal for malignancy or in male-to-female transgender individuals post-genital reassignment surgery which never felt their penises were part of their body image (26).

Transgender individuals may also experience distress regarding their genitalia, similar to the distress individuals with BID feel about the affected limb. Consistent with previous reports (27), there is comorbidity between BID and gender identity dysphoria, with 5 out of 18 individuals with BID in our sample exhibiting symptoms of both conditions. Our findings support the idea that both conditions may be associated with atypical activity of the rSPL; however, future studies comparing groups of individuals with BID and controls with and without gender identity dysphoria are needed for more clarity.

During observation of the desired body, hyperactivity in the rSPL was accompanied by corresponding hyperactivity in the rSMA. The rSMA is involved in initiating movements driven by internally generated plans rather than responses to external stimuli. It functions in motor planning, sequencing movements, and integrating sensory information relevant to motor actions (28, 29). It is also associated with the awareness and perception of self-generated movements, contributing to the sense of agency (30). The sense of agency involves efferent signals and continuous monitoring of body state, with the rSMA acting as a comparator (31). In BID, there may be a lapse in updating internal self-models during self-generated limb movement due to the lack of limb anchoring to the body image (32). Individuals with BID, unlike controls, report no differences in the perception of an actor’s body motion triggering their sensorimotor schema while pretending or being in a normal position (33). Also, our resting-state fMRI results support the framework for the missing anchoring of multimodal limb representation to body image. Before VR, we observed reduced functional connectivity between rAG/rSPL and visuo-occipital areas, including lCaudate, rCuneal, and rsLOC, aligning with previous findings of reduced structural connectivity in the parieto-occipital network for bodily self (7) indicating compromised visual processing of the affected limb and its integration into the body image (5, 8).

In experimental conditions where the undesired leg is visually faded (the disappearing limb trick (34)) or during pretending, alleviation of desire might occur by adjusting visual information to match the body image in the rSPL. VR offers an immersive method to resolve this discrepancy, normalizing altered parieto-occipital connectivity.

Reproducing prior results, functional connectivity alterations and subsequent normalization were observed in other brain networks, specifically the limbic and mirror motor networks, crucial for emotional processing and empathetic resonance with other mental states (35). Before VR, rAG exhibited reduced connectivity to rFP, which was also less connected to the PC. In the left hemisphere, lFP showed reduced connectivity to both lCaudate and laMTG, structures connected via the left uncinate fasciculus, a white matter tract altered in BID (7). Emotions are thought to be embodied processes shaped by physiological and social factors (36) and BID individuals, not surprisingly exhibit changes in the emotional domain due to the lack of body adherence (37). Alteration in the mirror system may lead to an overidentification with amputated body as observed by experimental data (18, 19) or autobiographic memories (38). Observing amputees triggers substantial resonance in the mirror motor network among healthy individuals, especially those with heightened empathy (39). Repetitive exposure to amputated bodies reduces the activity of this network in healthy individuals, excluding the occipital cortex and rSPL(40). In our study, embodying the desired body through VR in BID led to normalization of these neural networks.

Increased functional connectivity was observed in the reward system before VR, particularly in dopaminergic cortico-striatal tracts connecting the Accumbens to the IFGoper, lPreCG, and rSPL. Our findings align with the dopaminergic hypothesis in BID (23)-The activity of the comparator in the rSPL, and its possible deactivation through pretending but not embodied VR, may establish a reinforcing cycle increasing connectivity to the Accumbens. Phenomenological reports indicate that pretending can worsen intrusive thoughts in BID, similar to addiction (37). VR could serve as an effective and cost-efficient therapeutic method, restoring functional connectivity in the reward system.

In BID, the rIFG is hyperconnected to the nucleus accumbens. The rIFGoper is linked to limb disownership and misattribution in somatoparaphrenia, where right brain-damaged patients believe their contralesional limbs belong to someone else (41). While both conditions involve limb disownership, BID is developmental and non-delusional, without misattribution. In somatoparaphrenia, delusional disownership is due to a disconnection between the rIFG and the Broca area and left temporo-parietal junction (41). In BID, the rIFG-nucleus accumbens hyperconnection may reinforce disownership feelings without causing delusional misattribution. Normalizing this connectivity through VR may reduce the desire for amputation in BID patients, but may also have implications for somatoparaphrenia

This study aimed to replicate and strengthen the findings of previous research on structural alterations in BID (2, 4, 8). In a new, largely independent sample of BID individuals and controls, we found gray matter reductions in the same areas previously identified (8) (rSPL/rAG), as well as in regions related to mirror neurons (lMidFG, lPreCG) and the limbic and reward systems (lFOrb, lFP). Further discussion on the replication results, including picture ratings, is available in the SI.

Our study has several limitations. Firstly, the small sample size is due to the rarity and confidentiality of the disorder. Despite this, our sample of 18 participants is the largest in the current literature, achieved through years of personal contacts and financial support to facilitate global participation. Additionally, the clinical online data were not analyzed or discussed here due to a lack of control conditions, but we have shared the data and hope future researchers will expand this dataset.

Furthermore, we refrained from conducting correlation analyses in this study. This decision was made because the current dataset analysis was already extensive, and correlation analyses with a small sample size could yield misleading results.

Additionally, our task-based fMRI analysis did not reveal a three-way interaction (*Group* by *Session* by *Body State*), likely due to the limited number of participants and the design matrix containing eight blocks. Nevertheless, hypothesis-based contrasts yielded results in expected brain regions, even after whole-brain analyses with a conservative statistical threshold (p < 0.05 FWE voxel-wise corrected, k > 25).

In conclusion, the present study demonstrates the efficacy of VR as a therapeutic tool for BID, with the advantage of normalizing neurodivergent patterns in the bodily self, the mirror, and limbic system networks characteristic of the condition. Considering body image as innate raises questions about elective therapeutic approaches. Modifying an innate body image with a strong genetic component seems unrealistic. An alternative strategy could be the amputation which surveys suggest markedly increases quality-of-life scores (42). However, this option poses significant challenges, including post-operative treatment of phantom limb sensations (43). Pretending as described above, also has multiple adverse consequences.

A safer and more promising method appears to be VR which does not require direct supervision from medical or psychological professionals, can be performed at home, and is affordable. This study demonstrated that VR normalized altered patterns in the reward systems, suggesting it might be less addictive than pretending. Importantly, none of our participants reported an intensified desire for amputation after VR, in contrast to observations during pretending. An important consideration is the therapeutic plan, including the optimal frequency of exposure and whether regular VR practice could impact functional patterns and influence structural connectivity, leading to long-term stabilization of these effects(44)

Despite these unanswered questions, the positive phenomenological experiences reported by participants and the observed VR effects on the brain are encouraging. This study’s potential extends beyond BID, as VR methods could apply to various conditions affecting the bodily self. Ultimately, this study highlights how VR aids in understanding the seamless unity of body and self-experience that many take for granted.

## Acknowledgments

GS, YP, JH, BL were funded by the Swiss National Science Foundation (grant number: PP00P1_202674), and GS, BL, EC by the European Commission’s Horizon Europe Program through the INTELLIMAN project (https://intelliman-project.eu/, HORIZON-CL4-Digital-Emerging, grant number: 101070136). KR is funded by Health Research Board Ireland grant HRB-EIA-2019-003. We would like to thank Jannick Mauron and Noah Rischert for their valuable contributions as research assistants. Special thanks to the individuals with BID, who showed significant effort in traveling to Zurich from different locations of the world and committing to lengthy testing. We also would like to thank the control group for their participation.

## Author Contributions

Gianluca Saetta: Funding acquisition, Conceptualization, Methodology, Investigation, Data curation, Project administration, Formal analysis, Writing – Original draft; Yannik Peter: Investigation, Data curation, Project administration, Formal analysis, Writing – Reviewing and Editing. Kathy Ruddy: Supervision, Writing – Reviewing and Editing; Jasmine Ho:, Methodology, Writing – Reviewing and Editing; Roger Luechinger: Investigation, Data curation, Writing – Reviewing and Editing; Emily Cross: Supervision, Lars Michels: Methodology, Project administration, Supervision, Writing – Reviewing and Editing; Bigna Lenggenhager: Funding acquisition, Conceptualization Methodology, Project admini

## References

1. M. B. First, Desire for amputation of a limb: paraphilia, psychosis, or a new type of identity disorder. Psychol Med 35, 919–928 (2005).

2. R. M. Blom, et al., The Desire for Amputation or Paralyzation: Evidence for Structural Brain Anomalies in Body Integrity Identity Disorder (BIID). PLOS ONE 11, e0165789 (2016).

3. S. Fornaro, P. Patrikelis, G. Lucci, When having a limb means feeling overcomplete. Xenomelia, the chronic sense of disownership and the right parietal lobe hypothesis. Laterality 26, 564–583 (2021).

4. L. M. Hilti, et al., The desire for healthy limb amputation: structural brain correlates and clinical features of xenomelia. Brain 136, 318–329 (2013).

5. P. D. McGeoch, et al., Xenomelia: a new right parietal lobe syndrome. J. Neurol. Neurosurg. Psychiatr. 82, 1314–1319 (2011).

6. M. R. Longo, Body Image: Neural Basis of ‘Negative’ Phantom Limbs. Current Biology 30, R644–R646 (2020).

7. G. Saetta, et al., White matter abnormalities in the amputation variant of body integrity dysphoria. Cortex 151, 272–280 (2022).

8. G. Saetta, et al., Neural correlates of body integrity dysphoria. Current Biology 30, 2191–2195. e3 (2020).

9. P. Brugger, B. Lenggenhager, M. Giummarra, Xenomelia: A Social Neuroscience View of Altered Bodily Self-Consciousness. Frontiers in Psychology 4 (2013).

10. P. Brugger, M. Christen, L. Jellestad, J. Hänggi, Limb amputation and other disability desires as a medical condition. Lancet Psychiatry 3, 1176–1186 (2016).

11. M. B. First, C. E. Fisher, Body integrity identity disorder: the persistent desire to acquire a physical disability. Psychopathology 45, 3–14 (2012).

12. S. Chakraborty, G. Saetta, C. Simon, B. Lenggenhager, K. Ruddy, Could Brain–Computer Interface Be a New Therapeutic Approach for Body Integrity Dysphoria? Frontiers in Human Neuroscience 15, 433 (2021).

13. F. Scarpina, et al., The Effect of a Virtual-Reality Full-Body Illusion on Body Representation in Obesity. Journal of Clinical Medicine 8, 1330 (2019).

14. E. Bolt, et al., Effects of a virtual gender swap on social and temporal decision-making. Sci Rep 11, 15376 (2021).

15. P. Tacikowski, J. Fust, H. H. Ehrsson, Fluidity of gender identity induced by illusory body-sex change. Sci Rep 10, 14385 (2020).

16. C. Dijkerman, B. Lenggenhager, The body and cognition: The relation between body representations and higher level cognitive and social processes. Cortex 104, 133–139 (2018).

17. C. Turbyne, P. de Koning, J. Zantvoord, D. Denys, Body integrity identity disorder using augmented reality: a symptom reduction study. BMJ Case Reports CP 14, e238554 (2021).

18. J. T. Ho, G. Saetta, B. Lenggenhager, Influence of bodily states on cognition: A web-based study in individuals with body integrity dysphoria. Journal of Psychiatric Research 159, 66–75 (2023).

19. G. Macauda, R. Bekrater-Bodmann, P. Brugger, B. Lenggenhager, When less is more - Implicit preference for incomplete bodies in xenomelia. J Psychiatr Res 84, 249–255 (2017).

20. M. Scattolin, M. S. Panasiti, J. T. Ho, B. Lenggenhager, S. M. Aglioti, Ownership of the affected leg is further reduced following deceptive behaviors in body integrity dysphoria. iScience 26 (2023).

21. M. Tsakiris, G. Prabhu, P. Haggard, Having a body versus moving your body: How agency structures body-ownership. Consciousness and Cognition 15, 423–432 (2006).

22. M. Gandola, et al., Brain Abnormalities in Individuals with a Desire for a Healthy Limb Amputation: Somatosensory, Motoric or Both? A Task-Based fMRI Verdict. Brain Sci 11, 1248 (2021).

23. S. Oddo-Sommerfeld, et al., Brain activity elicited by viewing pictures of the own virtually amputated body predicts xenomelia. Neuropsychologia 108, 135–146 (2018).

24. P. Brugger, et al., Beyond re-membering: phantom sensations of congenitally absent limbs. Proc Natl Acad Sci U S A 97, 6167–6172 (2000).

25. P. D. McGeoch, V. S. Ramachandran, The appearance of new phantom fingers post-amputation in a phocomelus. Neurocase 18, 95–97 (2012).

26. P. D. McGeoch, V. S. Ramachandran, Phantom Phenomena—An Introduction to “Phantom Penis: Extrapolating Neuroscience and Employing Imagination for Trans Male Sexual Embodiment.” Studies in Gender and Sexuality 21, 247–250 (2020).

27. C. Scupin, T. Schnell, E. Kasten, How Defined Is Gender Identity in People with Body Integrity Dysphoria? Adv Mind Body Med 35, 17–32 (2021).

28. P. Haggard, Human volition: towards a neuroscience of will. Nat Rev Neurosci 9, 934–946 (2008).

29. L. Zapparoli, S. Seghezzi, E. Paulesu, The What, the When, and the Whether of Intentional Action in the Brain: A Meta-Analytical Review. Frontiers in Human Neuroscience 11 (2017).

30. P. Haggard, Sense of agency in the human brain. Nat Rev Neurosci 18, 196–207 (2017).

31. C. D. Frith, S.-J. Blakemore, D. M. Wolpert, Abnormalities in the awareness and control of action. Philosophical Transactions of the Royal Society of London. Series B: Biological Sciences 355, 1771–1788 (2000).

32. M. J. Giummarra, et al., Maladaptive Plasticity: Imprinting of Past Experiences Onto Phantom Limb Schemata. The Clinical Journal of Pain 27, 691–698 (2011).

33. G. Saetta, et al., Limb apparent motion perception: Modification by tDCS, and clinically or experimentally altered bodily states. Neuropsychologia 162, 108032 (2021).

34. K. D. Stone, F. Bullock, A. Keizer, H. C. Dijkerman, The disappearing limb trick and the role of sensory suggestibility in illusion experience. Neuropsychologia 117, 418–427 (2018).

35. F. Filimon, J. D. Nelson, D. J. Hagler, M. I. Sereno, Human cortical representations for reaching: Mirror neurons for execution, observation, and imagery. NeuroImage 37, 1315–1328 (2007).

36. L. W. Barsalou, Grounded cognition: past, present, and future. Top Cogn Sci 2, 716–724 (2010).

37. A. Pennisi, A. Capodici, “Bodies that love themselves and bodies that hate themselves: the role of lived experience in body integrity dysphoria” in Psychopathology and the Mind: What Mental Disorders Can Tell Us about Our Minds, (Routledge, 2021), pp. 222–252.

38. C. Obernolte, T. Schnell, E. Kasten, The role of specific experiences in childhood and youth in the development of body integrity identity disorder (BIID). American Journal of Applied Psychology 4, 1–8 (2015).

39. S.-L. Liew, T. Sheng, J. Margetis, L. Aziz-Zadeh, Both novelty and expertise increase action observation network activity. Frontiers in Human Neuroscience 7 (2013).

40. S.-L. Liew, T. Sheng, L. Aziz-Zadeh, Experience with an amputee modulates one’s own sensorimotor response during action observation. NeuroImage 69, 138–145 (2013).

41. G. Saetta, L. Michels, P. Brugger, Where in the Brain is “the Other’s” Hand? Mapping Dysfunctional Neural Networks in Somatoparaphrenia. Neuroscience 476, 21–33 (2021).

42. R. M. Blom, R. C. Hennekam, D. Denys, Body Integrity Identity Disorder. PLOS ONE 7, e34702 (2012).

43. S. Noll, E. Kasten, Body integrity identity disorder (BIID): how satisfied are successful wannabes. Psychology and Behavioral Sciences 3, 222–232 (2022).

44. A. May, Experience-dependent structural plasticity in the adult human brain. Trends in cognitive sciences 15, 475–482 (2011).

45. G. Stoet, PsyToolkit: A software package for programming psychological experiments using Linux. Behavior Research Methods 42, 1096–1104 (2010).

46. R. S. Desikan, et al., An automated labeling system for subdividing the human cerebral cortex on MRI scans into gyral based regions of interest. Neuroimage 31, 968–980 (2006).

47. A. Nieto-Castanon, Handbook of functional connectivity magnetic resonance imaging methods in CONN (Hilbert Press, 2020).

48. M. J. Jafri, G. D. Pearlson, M. Stevens, V. D. Calhoun, A Method for Functional Network Connectivity Among Spatially Independent Resting-State Components in Schizophrenia. Neuroimage 39, 1666–1681 (2008).

49. Y. Benjamini, Y. Hochberg, Controlling the False Discovery Rate: A Practical and Powerful Approach to Multiple Testing. Journal of the Royal Statistical Society. Series B (Methodological) 57, 289–300 (1995).

