## Supplemental materials for "Feeling at Home in a Virtually Amputated Body; Neural and Phenomenological Effects of Illusory Embodiment in Body Integrity Dysphoria"

### Supplementary Materials

#### Introduction – further hypotheses regarding replication of previous findings in an independent large BID group

Considering the early stage of BID research and the need to enhance our understanding of the condition, we also aimed to replicate previously observed brain structural alterations. Specifically, we sought to replicate prior findings indicating a reduction in gray matter concentration within specific brain regions associated with body image, multisensory integration, and the reward and limbic systems—namely the rSPL, the left premotor cortex, and orbitofrontal cortex( 1). In a largely independent cohort of individuals with BID. For additional replication, inspired by previous work (2), we integrated a task into our study design after the MRI sessions. In this task, participants were required to rate images, either of themselves or others, in both amputated and complete states. Consistent with their findings, we anticipated that individuals with BID would rate images of themselves in the amputated state as more emotionally satisfying, intense, attractive, and sexually arousing.

#### Task-based fMRI - MRI scanning and preprocessing steps

The Tesla Philips Ingenia whole-body scanner (Philips Medical Systems, Best, The Netherlands). had a transmit-receive body coil and a commercially available 32-element head coil array. For the task based fMRI protocol the Field of View (FOV) was 220mmx220mm, and the acquisition voxel size was 3.1mmx3.0mm. Thirty-five slices with a thickness of 3.0mm and a slice gap of 0.5mm were acquired. A sense factor of 1.5 was used. The scan duration consisted of 7 repetitions, each lasting 428 seconds. The repetition time and echo time were set at 2300ms and 35ms.
Data pre-processing and single-subject analysis were conducted using MATLAB (version 2016B) in conjunction with the SPM12 toolbox. A pre-processing pipeline was built up in R studio and the packages “spm12r” und “matlabr” was used to call SPM12 function in R studio. The script is available on OSF (https://osf.io/qag6u/). The pre-processing steps followed a standard procedure (3). Addressing motion-induced artifacts, motion realignment was performed, ensuring alignment across time points by registering functional MRI data to a reference image. To refine temporal alignment, slice-timing correction was applied. The anatomical reorientation and coregistration steps established anatomical-functional congruence. The anatomical MRI image was reoriented to a standardized orientation and subsequently coregistered to the mean functional MRI image. Subsequent anatomical MRI segmentation identified tissue types, while spatial normalization parameters were estimated to align the anatomical image to a common template. The data matrix was interpolated to yield 2 x 2 x 2 mm voxels. Further refining anatomical data, the segmentation isolated distinct tissue types, while spatial normalization, informed by the previous step, aligned the functional images. Spatial smoothing enhanced data quality by convolving functional MRI data with a 5x5x5 Gaussian filter, improving the signal-to-noise ratio.
Following preprocessing, the canonical hemodynamic response function (HRF) was employed to characterize the Blood-oxygen-level-dependent (BOLD) signal associated with each block relative to its baseline condition. This modeling enabled the identification of task-related neural activations. Artifacts originating from physiological noise, such as cardiac and respiratory cycles, were attenuated using high-pass filtering with a cutoff of 128 seconds. This filtering effectively removed low-frequency noise components from the data. To account for motion-related variability, the six motion parameters derived from each run were included as regressors in the analysis. This step aimed to control for any confounding effects introduced by head translations and rotations.

**RS-fMRI data - MRI scanning and preprocessing steps**

The rs-fMRI protocol involved a FOV of 221mmx221mm, with an acquisition voxel size of 2.9mmx3.0mm. 34 slices with a thickness of 3.0mm and a slice gap of 0.5mm were acquired. The sense factor for parallel imaging was set to 1.5. The scan duration was 496 seconds, and the repetition time and echo time values were 2300ms and 35ms. To minimize head motion, cushions were placed around the head. A full list of the parameters are reported in a separate data descriptor paper.

MATLAB (version 2016b, MathWorks, Natick, MA, USA) and CONN connectivity Toolbox version 22b were utilized for pre-processing and subsequent analyses. Functional and anatomical data were pre-processed using a flexible pre-processing pipeline (1) including realignment with correction of susceptibility distortion interactions, slice timing correction, outlier detection, direct segmentation and normalization (to the Montreal Neurological Institute (MNI) brain template), and spatial smoothing. Functional data were realigned using SPM realign & unwarp procedure, where all scans were coregistered to a reference image (first scan of the first session) using a least squares approach and a 6 parameter (rigid body) transformation and resampled using b-spline interpolation to correct for motion and magnetic susceptibility interactions. Temporal misalignment between different slices of the functional data was corrected following SPM slice-timing correction (STC) procedure, using sinc temporfal interpolation to resample each slice BOLD timeseries to a common mid-acquisition time. Potential outlier scans were identified using the artifact detection tool (ART) as acquisitions with framewise displacement above 0.9 mm or global BOLD signal changes above 5 standard deviations, and a reference BOLD image was computed for each subject by averaging all scans excluding outliers. Functional and anatomical data were normalized into standard MNI space, segmented into gray matter, white matter, and CSF tissue classes, and resampled to 2 mm isotropic voxels following a direct normalization procedure using SPM unified segmentation and normalization algorithm with the default IXI-549 tissue probability map template. Last, functional data were smoothed using spatial convolution with a Gaussian kernel of 8 mm full width half maximum.

In addition, functional data were denoised using a standard denoising pipeline including the regression of potential confounding effects characterized by white matter time series (5 CompCor noise components), CSF timeseries (5 CompCor noise components), and linear trends (2 factors) within each functional run. Next, a bandpass frequency filtering of the BOLD timeseries was applied between 0.008 Hz and 0.09 Hz. CompCor noise components within white matter and CSF were estimated by computing the average BOLD signal as well as the largest principal components orthogonal to the BOLD average within each subject's eroded segmentation masks.

**Supplementary Dataset for Replication Purposes**

**T1-weighted morphological data,**

The scanning procedure involved acquiring a high-resolution T1-weighted scan for each participant. For the 3D-T1 imaging protocol, the FOV was 240mmx240mm, with a high-resolution acquisition voxel size of 1x1mm. Thin slices of 1mm thickness were obtained, totalling 160 slices. The sense factor, indicating the level of parallel imaging, was set at 2.5. The scan duration for this sequence was 242 seconds, and the repetition time and echo time were 8.1ms and 3.7ms, respectively.

The Voxel-Based Morphometry (VBM) technique was utilized to detect variations in gray matter concentration patterns between BID and controls groups. The focus of the analysis

was primarily on the regions defined by the “BID VBM mask”, a mask created by merging the ROIs identified as atrophic or hypertrophic in individuals with BID in our preceding study (1). This mask is accessible on OSF (https://osf.io/qag6u/).
MATLAB R2016b and Statistical Parametric Mapping (SPM12. Wellcome Trust, UK) were employed for all data pre-processing and subsequent analyses. To ensure adherence to standardized procedures (1), the established pre-processing batch "preproc_vbm.m" from the SPM12 toolbox's "batches" directory was executed. Initially, image segmentation was conducted to distinguish between white and gray matter. Subsequently, DARTEL templates were generated to optimize the alignment of images. The transformation of gray matter into the MNI space occurred, and to address distortions introduced by stereotactic normalization, we applied Jacobian modulation.
Further processing included the normalization of Jacobian-scaled gray matter images using pre-estimated deformations. To address spatial variations, a Gaussian Kernel Gaussian Filter (8x8x8 $m^3$) was applied for spatial smoothing. The resulting smoothed images were averaged, and an explicit mask was derived by applying a threshold to the tissue average. The normalization process continued with bias-corrected images being aligned to the MNI space. Average normalized bias-corrected maps were then produced and assessed for consistency using SPM12's "Check Reg" tool.
Subsequent to pre-processing, the subject-specific gray matter maps were examined to identify anatomical differences between BID and control groups within anticipated key regions. This analysis involved a two-sample t-test for each voxel within the confines of the BID VBM mask.
To account for potential confounding factors, we opted for an ANCOVA approach for correction. This method allowed the identification of group differences in each voxel that were not solely explained by the linear relationship between voxel gray matter content and overall gray matter quantity for each participant. Results were extracted at $p < 0.01$ FWE voxel-wise corrected in line with the previous study (1). To improve visualisation, the results were displayed at $p < 0.001$ uncorrected.

**Task-related picture rating outside of the scanner**

Consistent with the approach taken in the study by Oddo-Sommerfeld and co-authors (2018), after the fMRI task and outside the scanner, participants were requested to assess the following attributes for each picture seen during the task-based fMRI using a VAS ranging from 0 (not at all) to 100 (very strong): *Pleasantness: “*How pleasant/unpleasant was the picture?*; Emotionality:* “How emotionally intense was the picture?*”* ; *Attractiveness*: “How appealing/attractive did the subjects in the pictures look?*”. Sexual Arousal: “*How sexually arousing was the picture*?”*.
The analysis was performed with same tools and applying the same parameter as described above (see virtual embodiment session). An ART-ANOVA examined the VAS score as a function of the between-factor Group (BID vs Control), the within-factors *Body State* (Full vs Amputated), *Person* (Self vs Other) and their interactions.

**Clinical data and online survey in BID**

Participants provided detailed information on their desire for limb amputation, including identification of targeted limb(s), emergence timeline, and subjective likelihood of amputation within a year (rated on a 0-20 scale, definitely not – definitely will have amputation). Responses, gathered online, covered various aspects such as intensity, social interaction avoidance, duration, relief through pretending, role in sexual fantasies, self-perception in dreams, body contentment, mood, emotional state, desire prominence, suffering

from BID symptoms, distress levels, and feelings of limb disownership. Questions also explored limb ownership and agency sensations.
Due to the absence of a control intervention group, involving individuals with BID undergoing either a control intervention or no intervention, the standalone data has not been extensively analysed due to its limited substantive relevance. Nevertheless, for comprehensive documentation and in a contribution to the scientific community, with the hope of ongoing data collection from other groups. tTe raw data is shared on OSF (https://osf.io/qag6u/).

#### Task-based fMRI supplementary results

Before to proceed with the contrast BID-PreVR-Amputated > BID-PostVR-Amputated, we inspected the effects of group and session. While there was no significant effect of Session, we found a significant effect of Group in the right fusiform gyrus, the left precentral gyrus, the left occipital fusiform gyrus, the right supplementary motor area and the right precuneus. Results are reported in the table below.

| **Brain regions** | **MNI coordinates** | | | | | | | |
|---|---|---|---|---|---|---|---|---|
| | **Left hemisphere** | | | | **Right hemisphere** | | | |
| | **x** | **y** | **z** | **Z-score** | **x** | **y** | **z** | **Z-score** |
| ***Main Effect of Group*** | | | | | | | | |
| Fusiform Gyrus | | | | | 38 | -46 | -16 | 7.58 |
| Precentral Gyrus | -44 | 2 | 32 | 6.62 | | | | |
| Occipital Fusiform Gyrus | -34 | -76 | -10 | 6.36 | | | | |
| Supplementary Motor Area | | | | | 6 | 12 | 50 | 5.74 |
| Precuneus | | | | | 12 | -66 | 38 | 5.67 |

**Table 1** Task-based fMRI results. Main effect of Group

#### Resting-state fMRI supplementary results

Also for the resting-state fMRI analysis before to inspect the contrast Control > BID, PreVR > PostVR we looked at the main effect of Group (results displayed in Figure + and Session. Results are displayed below.

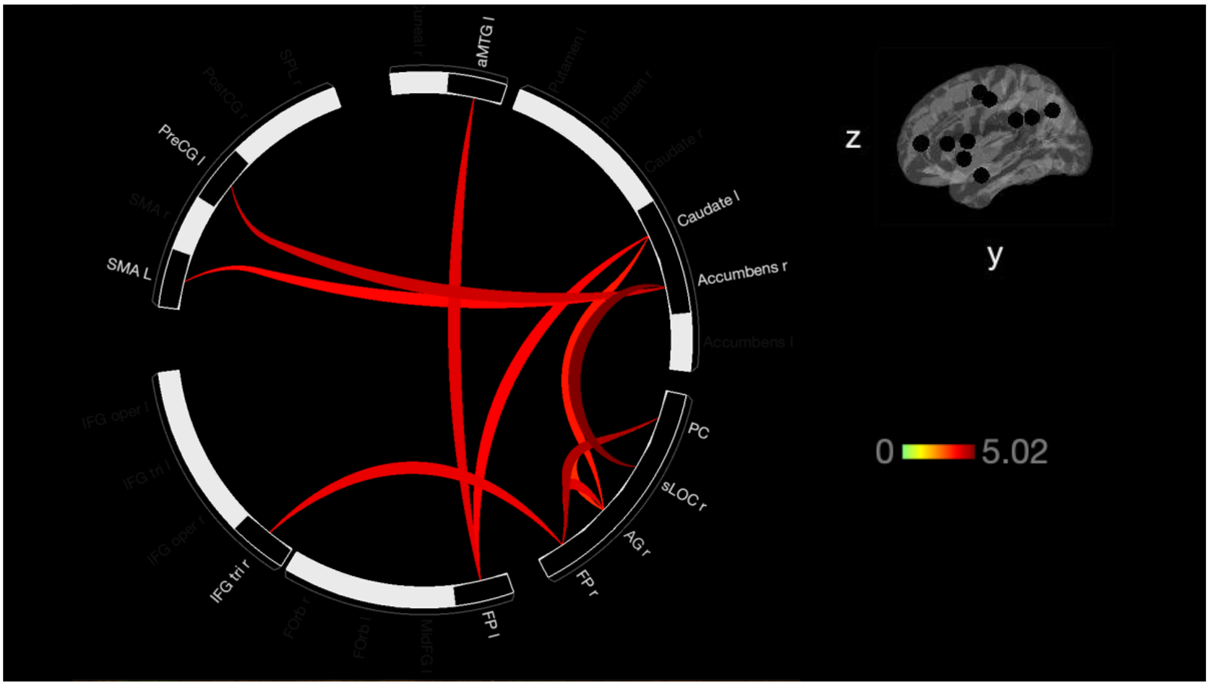

**Figure 1 SOM.** Resting-state fMRI results. Main effect of group Acronymous specified in the data-analysis section.

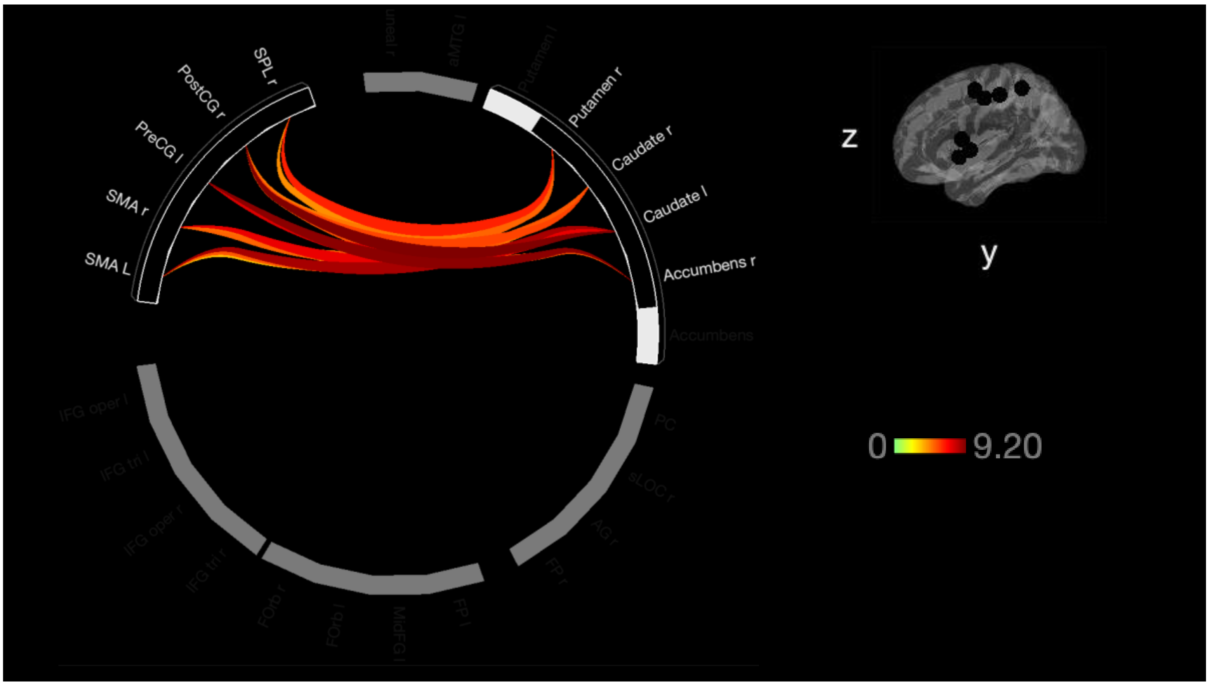

**Figure 2 SOM**. Resting-state fMRI results. Main effect of Session. Acronymous specified in the data-analysis section.

Task-related picture rating outside of the scanner supplementary results

We hypothesised that individuals with BID would rate the pictures with amputation as more emotionally gratifying, intense, attractive, and sexually arousing compared to controls.

For *Pleasantness*, the ART-ANOVA revealed a statistically significant effect of *Group* ($F = 20.43$, $p < 0.001$). The factor *Body State* also showed a statistically significant effect (F =

11.72, p < 0.001). The factor *Person* demonstrated a significant effect as well (F = 8.94, p < 0.001). The two-way interaction effect between *Group* and *Body State* was found to be highly significant (F = 173.91789, p < 0.001). Additionally, the three-way interaction effect between *Group*, *Body State*, and *Person* was also significant (F = 27.94, p < 0.001).
For *Emotional Intensity*, a main effect was seen for the factors *Group* (F = 10.30, p < 0.001), *Body State* (F = 100.36, p < 0.001), and *Person* (F = 54.90, p < 0.001). The two-way interaction effect between *Group* and *Body State* was significant (F = 82.92, p < 0.001). Additionally, the interaction effect between *Group* and *Person* was significant (F = 5.27, p < 0.001), and the interaction effect between *Body State* and *Person* was significant (F = 4.44, p = 0.036). The three-way interaction effect between *Group*, *Body State*, and *Person* was also significant (F = 7.3354, p = 0.007).
For *Appeal/Attractiveness*, the ART-ANOVA revealed a main effect of *Group* (F = 6.17, p = 0.018) and *Person* (F = 24.25, p < 0.001), no main effect of *Body State* (F = 2.89, p = 0.09). The two-way interaction effect between *Group* and *Body State* was found to be highly significant (F = 49.79, p < 0.001). The two-way interaction between *Group* and *Person*, and between *Body State* and *Person* were not significant Additionally, the three-way interaction effect between *Group, Body State, and Person* was also significant (F = 6.23, p = 0.013).
For *Sexual Arousal*, a main effect was observed for factors *Group* (F = 7.36, p = 0.01), *Body State* (F = 41.76, p < 0.001), and *Person* (F = 31.70, p < 0.001). The two-way interaction effect between *Group* and *Body State* was highly significant (F = 88.82, p < 0.001). Additionally, the interaction effect between *Group* and *Person* was significant (F = 21.48, p < 0.001), as well as the interaction effect between *Body State* and *Person* (F = 7.69, p = 0.005). The three-way interaction effect between *Group*, *Body State*, and *Person* was also significant (F = 19.70, p < 0.001).
Results are shown in Fig. 3 SOM. Post-hoc results are provided at the end of the SOM. Scripts and data are shared on OSF (https://osf.io/qag6u/).

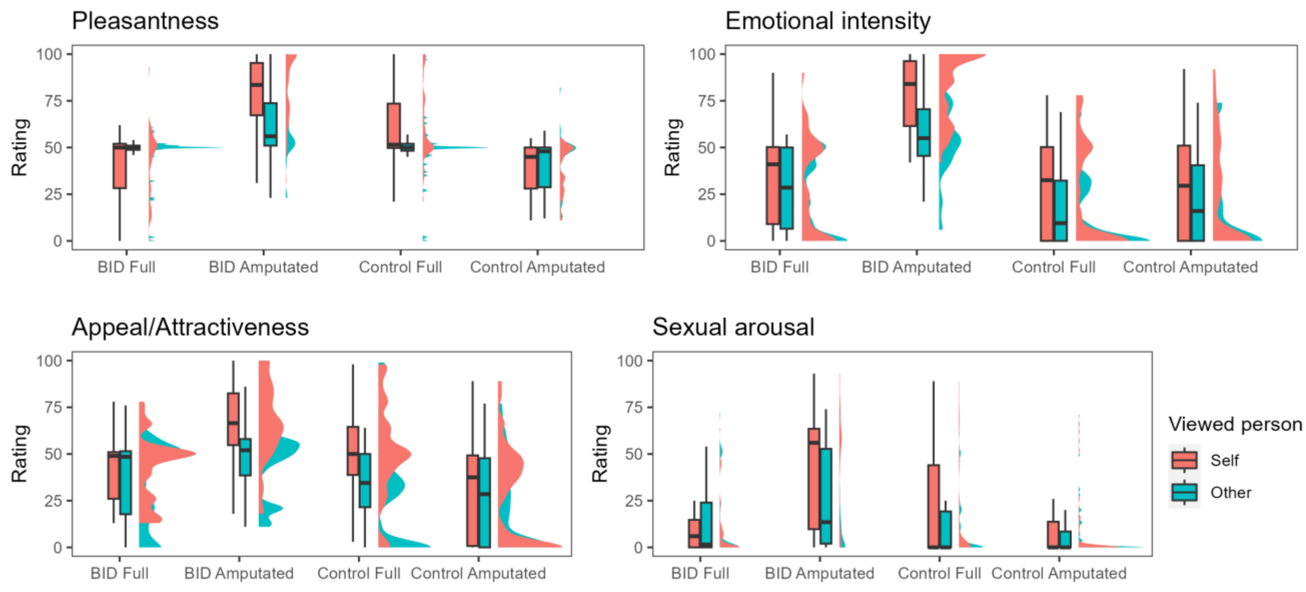


**Figure 3 SOM**. Results of the picture rating task. Boxplots with medians and interquartile ranges are presented.

#### Supplementary discussion

Another objective of the current study was to replicate and enhance the robustness of previously identified structural alterations in BID (1, 4, 5). In a distinct and largely independent sample of individuals with BID compared to controls, we observed gray matter reductions in precisely the same areas identified in Saetta et al., (2020), namely rSPL/rAG, as well as in other regions associated with mirror neurons (i.e., the lMidFG, the lPreCG) and regions of the limbic and reward systems (the lFOrb, the lFP). Further discussion for replication results including those about picture ratings founds place in the SOM This finding is noteworthy, considering that unlike the previous study, which exclusively recruited participants with a left leg amputation, the present study included individuals with right or left leg amputation desire. This difference may explain why we did not find functional connectivity or activity alterations in the right paracentral lobule, which is the region associated with the sensorimotor representation of the left leg. However, these results provide valuable information, suggesting that the engagement of the same brain region in BID, particularly the rSPL, may not significantly depend on the laterality of the affected leg. Additionally, replicating the findings of Oddo-Sommerfeld et al. (2018), we showed that Individuals with BID consistently rated pictures depicting their desired body state (that of an amputee) as more pleasant, intense, attractive, and sexually arousing compared to pictures of their undesired body state (real body). In contrast, control subjects rated pictures of their desired body state (real body) as more pleasant, intense, and attractive, without the heightened sexual arousal associated with the amputee body state. The association between the desire for amputation, the mental imagery of an amputee, or observing an amputee, and sexual arousal in most individuals with BID is a well-documented phenomenon (6, 7). Additionally, existing literature suggests that the body image in the brain can influence sexual preferences for bodies aligned with the individual's body image (8). Also, the clear double dissociation between BID and control groups underscores the distinct preferences of the two groups for markedly different body states. The presence of such a double dissociation may open the possibility for the development of a new neuropsychological clinical test to differentiate between individuals with BID and controls based on these ratings. Future validation of such neuropsychological tests is warranted.

**References**


1. G. Saetta, *et al.*, Neural correlates of body integrity dysphoria. *Curr. Biol.* 30, 2191-2195. e3 (2020).

2. S. Oddo-Sommerfeld, *et al.*, Brain activity elicited by viewing pictures of the own virtually amputated body predicts xenomelia. *Neuropsychologia* 108, 135–146 (2018).

3. F. Morfini, S. Whitfield-Gabrieli, A. Nieto-Castañón, Functional connectivity MRI quality control procedures in CONN. *Front. Neurosci.* 17, 1092125 (2023).

4. R. M. Blom, *et al.*, The Desire for Amputation or Paralyzation: Evidence for Structural Brain Anomalies in Body Integrity Identity Disorder (BIID). *PLOS ONE* 11, e0165789 (2016).

5. L. M. Hilti, *et al.*, The desire for healthy limb amputation: structural brain correlates and clinical features of xenomelia. *Brain* 136, 318–329 (2013).

6. R. M. Blom, S. J. van der Wal, N. C. Vulink, D. Denys, Role of Sexuality in Body Integrity Identity Disorder (BIID): A Cross-Sectional Internet-Based Survey Study. *J. Sex. Med.* 14, 1028–1035 (2017).

7. P. Brugger, M. Christen, L. Jellestad, J. Hänggi, Limb amputation and other disability desires as a medical condition. *Lancet Psychiatry* 3, 1176–1186 (2016).

8. V. S. Ramachandran, D. Brang, P. D. McGeoch, W. Rosar, Sexual and food preference in apotemnophilia and anorexia: interactions between "beliefs" and "needs" regulated by two-way connections between body image and limbic structures. *Perception* 38, 775–777 (2009).

**Questionnaire on Virtual Embodiment**

Participants completed a seven-item questionnaire designed to assess various aspects of embodiment using a visual analogue scale (VAS) graded from 0 to 100 (0 = not at all, 100 = very strong). The exact questions and their corresponding conditions are listed below:

1. Affected Leg Disownership:
    - "How strong is the feeling that the affected leg is not yours?"
2. Virtual Body Ownership:
    - "How strong is the feeling that the virtual body is your body?"
3. Virtual Body Agency:
    - "How strong is the feeling that the movements of the virtual body are your movements?"
4. Virtual Leg Ownership:
    - "How strong is the feeling that the left leg is your left leg?"
    - "How strong is the feeling that the right leg is your right leg?"
5. Virtual Leg Agency:
    - "How strong is the feeling that the movements of the virtual left leg are your movements?"
    - "How strong is the feeling that the movements of the virtual right leg are your movements?"

Participants used head movements to control a cursor, indicating their responses on a displayed bar without seeing the selected value.

**Online Survey Questions and Complementary Inquiries to the Zurich Xenomelia Scale**

- Which limb(s) on which side of your body are affected by your BID symptoms?
- Which of your two legs would you amputate first if you had to choose? If no answer is possible we will randomly assign one of the legs.
- Have you ever had an amputation in the past?
- At what age did your BID symptoms first appear?
- How do you usually see yourself in your dreams? with intact limb(s)
- In your opinion, what is the likelihood that you will undergo amputation of your unwanted limb(s) within the next year?
- My desire for amputation is so strong that it determines my life
- I have never played with the thought to amputate myself/to provoke an accident
- If I could choose between a sexual partner with an amputation and one without (everything else equal), I would go for the one without amputation
- I am far from moving and behaving as if I were amputated
- Despite the fact that I would have a body part removed, I would feel more “complete” after the desired amputation
- If I were amputated, I would experience myself as more erotic
- Instruments commonly used by amputees (prostheses, crutches, calipers, wheelchairs a.o.) do not fascinate me in any way
- I sometimes pretend (for myself or for others) to be amputated
- The theme of amputation plays an important role in my erotic fantasies
- However present, my desire for amputation is probably rather playful, i.e., a not-so-serious fantasy
- If I succeeded to make people around me believe that I am already amputated, it could reduce my desire for actual amputation
- For myself, the desire for amputation does not have any erotic or sexual connotation
- How strong was your desire for amputation of the affected limb(s) last week?
- To what extent did you avoid social contact last week because of your BID symptoms?
- During the past week, how many hours did you spend simulating your desired body condition (e.g., by bandaging your legs, in a wheelchair, or using crutches)?
- How much did such simulating behaviors relieve your symptoms?
- How satisfied were you with your body last week?
- How much did you suffer from your BID symptoms last week?
- How much was your general discomfort regarding your BID symptoms last week?
- How strong was your feeling of alienation for your unwanted limb(s) last week?

- What was your overall mood last week?
- How did you feel during the past week?
- How tired/awake were you during the past week?
- How tense/relaxed were you during the past week?
- How sexually aroused are you by the thought of being an amputee?
- How strongly do you feel that the following limbs are part of your body? left arm
- How strongly do you feel that the following limbs are part of your body? right arm
- How strongly do you feel that the following limbs are part of your body? left leg
- How strongly do you feel that the following limbs are part of your body? right leg
- How strongly do you feel that the movements of the following limbs are controlled by you? left arm
- How strongly do you feel that the movements of the following limbs are controlled by you? right arm
- How strongly do you feel that the movements of the following limbs are controlled by you? left leg
- How strongly do you feel that the movements of the following limbs are controlled by you? right leg

#### Task-related picture rating outside of the scanner – Post-hoc tests

**Table 1. Pleasantness**

*Significant interaction Group by Body State by Person. Post-hoc test comparisons*

| contrast | estimate | *SE* | *df* | *t* | *p* |
|---|---|---|---|---|---|
| bid,amputated,Other - bid,amputated,Self | -45.71 | 13.94 | 246 | -3.28 | .026* |
| bid,amputated,Other - bid,full,Other | 69.71 | 13.94 | 246 | 5.00 | < .001*** |
| bid,amputated,Other - bid,full,Self | 84.07 | 13.94 | 246 | 6.03 | < .001*** |
| bid,amputated,Other - control,amputated,Other | 76.69 | 23.12 | 246 | 3.32 | .023* |
| bid,amputated,Other - control,amputated,Self | 87.99 | 23.12 | 246 | 3.81 | .004** |
| bid,amputated,Other - control,full,Other | 38.94 | 12.08 | 246 | 3.23 | .031* |
| bid,amputated,Other - control,full,Self | -0.14 | 23.12 | 246 | -0.01 | 1.00 |
| bid,amputated,Self - bid,full,Other | 115.42 | 13.94 | 246 | 8.28 | < .001*** |
| bid,amputated,Self - bid,full,Self | 129.78 | 13.94 | 246 | 9.31 | < .001*** |
| bid,amputated,Self - control,amputated,Other | 122.40 | 23.12 | 246 | 5.29 | < .001*** |
| bid,amputated,Self - control,amputated,Self | 133.69 | 23.12 | 246 | 5.78 | < .001*** |
| bid,amputated,Self - control,full,Other | 84.65 | 12.08 | 246 | 7.01 | < .001*** |
| bid,amputated,Self - control,full,Self | 45.57 | 23.12 | 246 | 1.97 | .504 |
| bid,full,Other - bid,full,Self | 14.36 | 13.94 | 246 | 1.03 | .969 |
| bid,full,Other - control,amputated,Other | 6.99 | 23.12 | 246 | 0.30 | 1.00 |
| bid,full,Other - control,amputated,Self | 18.28 | 23.12 | 246 | 0.79 | .993 |
| bid,full,Other - control,full,Other | -30.76 | 12.08 | 246 | -2.55 | .181 |
| bid,full,Other - control,full,Self | -69.85 | 23.12 | 246 | -3.02 | .055 |
| bid,full,Self - control,amputated,Other | -7.38 | 23.12 | 246 | -0.32 | 1.00 |
| bid,full,Self - control,amputated,Self | 3.92 | 23.12 | 246 | 0.17 | 1.00 |
| bid,full,Self - control,full,Other | -45.12 | 12.08 | 246 | -3.74 | .006** |
| bid,full,Self - control,full,Self | -84.21 | 23.12 | 246 | -3.64 | .008** |
| control,amputated,Other - control,amputated,Self | 11.29 | 13.94 | 246 | 0.81 | .992 |
| control,amputated,Other - control,full,Other | -37.75 | 13.94 | 246 | -2.71 | .125 |
| control,amputated,Other - control,full,Self | -76.83 | 13.94 | 246 | -5.51 | < .001*** |
| control,amputated,Self - control,full,Other | -49.04 | 13.94 | 246 | -3.52 | .012* |
| control,amputated,Self - control,full,Self | -88.12 | 13.94 | 246 | -6.32 | < .001*** |
| control,full,Other - control,full,Self | -39.08 | 13.94 | 246 | -2.80 | .099 |

*Note.* * p < .05, ** p < .01, *** p < .001

**Table 2. Emotional Intensity**

*Significant interaction Group by Body State by Person. Post-hoc test comparisons*

| contrast | estimate | *SE* | *df* | *t* | *p* |
|---|---|---|---|---|---|
| bid,amputated,Other - bid,amputated,Self | -45.71 | 13.94 | 246 | -3.28 | .026* |
| bid,amputated,Other - bid,full,Other | 69.71 | 13.94 | 246 | 5.00 | < .001*** |
| bid,amputated,Other - bid,full,Self | 84.07 | 13.94 | 246 | 6.03 | < .001*** |
| bid,amputated,Other - control,amputated,Other | 76.69 | 23.12 | 246 | 3.32 | .023* |
| bid,amputated,Other - control,amputated,Self | 87.99 | 23.12 | 246 | 3.81 | .004** |
| bid,amputated,Other - control,full,Other | 38.94 | 12.08 | 246 | 3.23 | .031* |
| bid,amputated,Other - control,full,Self | -0.14 | 23.12 | 246 | -0.01 | 1.00 |
| bid,amputated,Self - bid,full,Other | 115.42 | 13.94 | 246 | 8.28 | < .001*** |
| bid,amputated,Self - bid,full,Self | 129.78 | 13.94 | 246 | 9.31 | < .001*** |
| bid,amputated,Self - control,amputated,Other | 122.40 | 23.12 | 246 | 5.29 | < .001*** |
| bid,amputated,Self - control,amputated,Self | 133.69 | 23.12 | 246 | 5.78 | < .001*** |
| bid,amputated,Self - control,full,Other | 84.65 | 12.08 | 246 | 7.01 | < .001*** |

**Table 2. Emotional Intensity**

*Significant interaction Group by Body State by Person. Post-hoc test comparisons*

| contrast | estimate | *SE* | *df* | *t* | *p* |
|---|---|---|---|---|---|
| bid,amputated,Self - control,full,Self | 45.57 | 23.12 | 246 | 1.97 | .504 |
| bid,full,Other - bid,full,Self | 14.36 | 13.94 | 246 | 1.03 | .969 |
| bid,full,Other - control,amputated,Other | 6.99 | 23.12 | 246 | 0.30 | 1.00 |
| bid,full,Other - control,amputated,Self | 18.28 | 23.12 | 246 | 0.79 | .993 |
| bid,full,Other - control,full,Other | -30.76 | 12.08 | 246 | -2.55 | .181 |
| bid,full,Other - control,full,Self | -69.85 | 23.12 | 246 | -3.02 | .055 |
| bid,full,Self - control,amputated,Other | -7.38 | 23.12 | 246 | -0.32 | 1.00 |
| bid,full,Self - control,amputated,Self | 3.92 | 23.12 | 246 | 0.17 | 1.00 |
| bid,full,Self - control,full,Other | -45.12 | 12.08 | 246 | -3.74 | .006** |
| bid,full,Self - control,full,Self | -84.21 | 23.12 | 246 | -3.64 | .008** |
| control,amputated,Other - control,amputated,Self | 11.29 | 13.94 | 246 | 0.81 | .992 |
| control,amputated,Other - control,full,Other | -37.75 | 13.94 | 246 | -2.71 | .125 |
| control,amputated,Other - control,full,Self | -76.83 | 13.94 | 246 | -5.51 | < .001*** |
| control,amputated,Self - control,full,Other | -49.04 | 13.94 | 246 | -3.52 | .012* |
| control,amputated,Self - control,full,Self | -88.12 | 13.94 | 246 | -6.32 | < .001*** |
| control,full,Other - control,full,Self | -39.08 | 13.94 | 246 | -2.80 | .099 |

*Note.* * p < .05, ** p < .01, *** p < .001

**Table 3. Appeal/Attractiveness**

*Significant interaction Group by Body State by Person. Post-hoc test comparisons*

| contrast | estimate | *SE* | *df* | *t* | *p* |
|---|---|---|---|---|---|
| bid,amputated,Other - bid,amputated,Self | -52.26 | 14.13 | 246 | -3.70 | .006** |
| bid,amputated,Other - bid,full,Other | 44.39 | 14.13 | 246 | 3.14 | .039* |
| bid,amputated,Other - bid,full,Self | 29.69 | 14.13 | 246 | 2.10 | .416 |
| bid,amputated,Other - control,amputated,Other | 15.50 | 23.42 | 246 | 0.66 | .998 |
| bid,amputated,Other - control,amputated,Self | 2.76 | 23.42 | 246 | 0.12 | 1.00 |
| bid,amputated,Other - control,full,Other | -1.88 | 12.23 | 246 | -0.15 | 1.00 |
| bid,amputated,Other - control,full,Self | -53.21 | 23.42 | 246 | -2.27 | .314 |
| bid,amputated,Self - bid,full,Other | 96.65 | 14.13 | 246 | 6.84 | < .001*** |
| bid,amputated,Self - bid,full,Self | 81.96 | 14.13 | 246 | 5.80 | < .001*** |
| bid,amputated,Self - control,amputated,Other | 67.76 | 23.42 | 246 | 2.89 | .078 |
| bid,amputated,Self - control,amputated,Self | 55.03 | 23.42 | 246 | 2.35 | .272 |
| bid,amputated,Self - control,full,Other | 50.39 | 12.23 | 246 | 4.12 | .001** |
| bid,amputated,Self - control,full,Self | -0.94 | 23.42 | 246 | -0.04 | 1.00 |
| bid,full,Other - bid,full,Self | -14.69 | 14.13 | 246 | -1.04 | .968 |
| bid,full,Other - control,amputated,Other | -28.89 | 23.42 | 246 | -1.23 | .921 |
| bid,full,Other - control,amputated,Self | -41.63 | 23.42 | 246 | -1.78 | .636 |
| bid,full,Other - control,full,Other | -46.26 | 12.23 | 246 | -3.78 | .005** |
| bid,full,Other - control,full,Self | -97.60 | 23.42 | 246 | -4.17 | .001** |
| bid,full,Self - control,amputated,Other | -14.19 | 23.42 | 246 | -0.61 | .999 |
| bid,full,Self - control,amputated,Self | -26.93 | 23.42 | 246 | -1.15 | .945 |
| bid,full,Self - control,full,Other | -31.57 | 12.23 | 246 | -2.58 | .168 |
| bid,full,Self - control,full,Self | -82.90 | 23.42 | 246 | -3.54 | .011* |
| control,amputated,Other - control,amputated,Self | -12.74 | 14.13 | 246 | -0.90 | .986 |
| control,amputated,Other - control,full,Other | -17.37 | 14.13 | 246 | -1.23 | .922 |
| control,amputated,Other - control,full,Self | -68.71 | 14.13 | 246 | -4.86 | < .001*** |
| control,amputated,Self - control,full,Other | -4.64 | 14.13 | 246 | -0.33 | 1.00 |
| control,amputated,Self - control,full,Self | -55.97 | 14.13 | 246 | -3.96 | .002** |
| control,full,Other - control,full,Self | -51.33 | 14.13 | 246 | -3.63 | .008** |

*Note.* * p < .05, ** p < .01, *** p < .001

**Table 4. Sexual arousal**

*Significant interaction Group by Body State by Person. Post-hoc test comparisons*

| contrast | estimate | *SE* | *df* | *t* | *p* |
|---|---|---|---|---|---|
| bid,amputated,Other - bid,amputated,Self | -52.26 | 14.13 | 246 | -3.70 | .006** |
| bid,amputated,Other - bid,full,Other | 44.39 | 14.13 | 246 | 3.14 | .039* |
| bid,amputated,Other - bid,full,Self | 29.69 | 14.13 | 246 | 2.10 | .416 |
| bid,amputated,Other - control,amputated,Other | 15.50 | 23.42 | 246 | 0.66 | .998 |
| bid,amputated,Other - control,amputated,Self | 2.76 | 23.42 | 246 | 0.12 | 1.00 |
| bid,amputated,Other - control,full,Other | -1.88 | 12.23 | 246 | -0.15 | 1.00 |
| bid,amputated,Other - control,full,Self | -53.21 | 23.42 | 246 | -2.27 | .314 |
| bid,amputated,Self - bid,full,Other | 96.65 | 14.13 | 246 | 6.84 | < .001*** |
| bid,amputated,Self - bid,full,Self | 81.96 | 14.13 | 246 | 5.80 | < .001*** |

**Table 4. Sexual arousal**

*Significant interaction Group by Body State by Person. Post-hoc test comparisons*

| contrast | estimate | *SE* | *df* | *t* | *p* |
|---|---|---|---|---|---|
| bid,amputated,Self - control,amputated,Other | 67.76 | 23.42 | 246 | 2.89 | .078 |
| bid,amputated,Self - control,amputated,Self | 55.03 | 23.42 | 246 | 2.35 | .272 |
| bid,amputated,Self - control,full,Other | 50.39 | 12.23 | 246 | 4.12 | .001** |
| bid,amputated,Self - control,full,Self | -0.94 | 23.42 | 246 | -0.04 | 1.00 |
| bid,full,Other - bid,full,Self | -14.69 | 14.13 | 246 | -1.04 | .968 |
| bid,full,Other - control,amputated,Other | -28.89 | 23.42 | 246 | -1.23 | .921 |
| bid,full,Other - control,amputated,Self | -41.63 | 23.42 | 246 | -1.78 | .636 |
| bid,full,Other - control,full,Other | -46.26 | 12.23 | 246 | -3.78 | .005** |
| bid,full,Other - control,full,Self | -97.60 | 23.42 | 246 | -4.17 | .001** |
| bid,full,Self - control,amputated,Other | -14.19 | 23.42 | 246 | -0.61 | .999 |
| bid,full,Self - control,amputated,Self | -26.93 | 23.42 | 246 | -1.15 | .945 |
| bid,full,Self - control,full,Other | -31.57 | 12.23 | 246 | -2.58 | .168 |
| bid,full,Self - control,full,Self | -82.90 | 23.42 | 246 | -3.54 | .011* |
| control,amputated,Other - control,amputated,Self | -12.74 | 14.13 | 246 | -0.90 | .986 |
| control,amputated,Other - control,full,Other | -17.37 | 14.13 | 246 | -1.23 | .922 |
| control,amputated,Other - control,full,Self | -68.71 | 14.13 | 246 | -4.86 | < .001*** |
| control,amputated,Self - control,full,Other | -4.64 | 14.13 | 246 | -0.33 | 1.00 |
| control,amputated,Self - control,full,Self | -55.97 | 14.13 | 246 | -3.96 | .002** |
| control,full,Other - control,full,Self | -51.33 | 14.13 | 246 | -3.63 | .008** |

*Note.* * p < .05, ** p < .01, *** p < .001
